# Divergent patterns of genetic overlap between severe mental disorders and metabolic markers

**DOI:** 10.1101/2024.11.04.24316693

**Authors:** Dennis van der Meer, Alexey A. Shadrin, Sara E. Stinson, Elise Koch, Jaroslav Rokicki, Zillur Rahman, Jacob Bergstedt, Aigar Ottas, Ida E. Sønderby, Linn Rødevand, Julian Fuhrer, Daniel S. Quintana, Anders M. Dale, Kevin S. O’Connell, Srdjan Djurovic, Kelli Lehto, Maris Alver, Lili Milani, Ole A. Andreassen

## Abstract

**Background:** Bipolar disorder (BIP), major depressive disorder (MDD) and schizophrenia (SCZ) are severe mental disorders (SMDs), each associated with poor cardiometabolic health. Mapping the genetic relationships of these highly heritable disorders with blood markers of metabolic activity may uncover biological pathways underlying this important shared clinical feature.

**Methods:** We charted global genetic overlap of the three SMDs, type 2 diabetes (T2D), coronary artery disease (CAD), and body mass index (BMI) with 249 circulating metabolic markers through linkage disequilibrium score regression and bivariate Gaussian mixture modeling. We estimated causal relationships, functionally annotated shared genetic variants, and investigated enrichment across diverse brain and body tissues.

**Results:** All three SMDs had extensive overlap with the metabolic markers. The pattern of genetic correlations was highly similar between MDD, T2D, CAD, and BMI (Spearman’s correlation rs>.93), opposite in direction to the pattern found for SCZ and BIP (MDD-BIP rs=-.74; MDD-SCZ rs=-.83). The metabolic markers had widespread, robust causal effects on the SMDs and cardiometabolic traits. We mapped 1056 genes shared between the individual SMDs and the metabolic markers to disorder-specific processes related to metabolic activity, mitochondrial function, and synaptic processes. These genes were most prominently expressed throughout the brain, heart and liver.

**Discussion:** SMDs have strong associations with metabolic markers, whereby MDD has a distinctly different genetic relationship than BIP and SCZ. Our findings suggest that metabolic pathways are involved in the development of SMDs and can play a central role in disentangling disorder-specific etiologies. Our “metabolic psychiatry” approach has high potential to guide the development of targeted interventions.

---

Bipolar disorder (BIP), major depressive disorder (MDD), and schizophrenia (SCZ) are severe mental disorders (SMDs) with shared etiology and overlapping clinical features.^1^ These disorders are also associatied with higher rates of cardiometabolic diseases such as type 2 diabetes (T2D) and coronary artery disease (CAD), which significantly impact clinical outcomes, quality of life and life expectancy.^2^ Further, dyslipidemia and weight gain are common adverse side effects of psychotropic medication,^3^ which are linked to treatment response and non-adherence.^4,5^ SMDs and cardiometabolic diseases rank first and second in the global burden of disease.^6^ Thus, uncovering their shared determinants may be highly impactful for improving public health globally.

Metabolic processes produce sets of metabolites that may act as fingerprints, capturing their activity. Medication-naïve individuals with SCZ as well as their siblings have abnormal levels of a range of these markers, indicating defects in lipid metabolism and glycolysis.^7^ Such dysfunction has also been reported for BIP and MDD.^8,9^ Panels of markers can thereby form a biological signature that separates individuals with SMDs from controls.^10^ Markers such as lipoproteins, cholesterol, fatty acids, and glucose are now well-captured in plasma by high-throughput nuclear magnetic resonance (NMR) spectroscopy,^11^ explaining more variance in metabolic dysfunction than traditional measurements.^12^

Recent genome-wide association studies (GWAS) of SMDs,^13–15^ cardiometabolic diseases^16,17^ and metabolic markers^18^ have quantified the genetic architectures of these phenotypes, and discovered hundreds of associated loci. The resulting well-powered GWAS summary statistics, combined with a new generation of biostatistical tools,^19^ enable us to accurately map genetic overlap and estimate causal relationships, less burdened by the influence of secondary disease processes and medication use that may confound estimates of phenotypic associations.

Here, we chart the genetic relationships between SMDs and metabolic markers, with the overall goal to better understand the connection between mental and cardiometabolic health. Specifically, we aim to characterize and compare the global and local genetic overlap of SCZ, BIP and MDD with metabolic markers, and determine underlying pathways and body-brain relationships. Identification of the biological pathways involved could pave the way towards novel therapeutic strategies, ultimately improving the diagnosis, treatment, and prevention of SMDs and comorbid cardiometabolic diseases.

## Results

### Global genetic overlap

For the analyses of genetic overlap, we included GWAS summary statistics of the Nightingale panel of metabolic markers,^11^ encompassing 228 lipids, lipoproteins or fatty acids and 21 non-lipid traits, namely amino acids, ketone bodies, fluid balance, glycolysis-, and inflammation-related metabolic markers,^18^ see Supplementary Table 1 for an overview of these markers, with abbreviations and categorizations. We further used the latest and most powerful GWAS of BIP,^15^ MDD,^14^ and SCZ,^13^ as well as T2D,^17^ and CAD,^16^ as commonly comorbid cardiometabolic conditions. We also included body mass index (BMI, as proxy of obesity),^20^ a known risk factor of these conditions. To correct for comparisons across the 249 markers, we adjusted p-values with the Benjamini and Hochberg method, and set significance thresholds at α=.05.

First, we calculated the genetic correlation between each of the metabolic markers and the included traits, using LD Score Regression (LDSC).^21^ This revealed extensive genetic relationships of the markers with all three SMDs, though the extent varied across disorders. MDD had multiple comparisons-corrected significant genetic correlations with 194 of 249 markers, compared to 109 for SCZ and 7 for BIP. **Figure 1a** summarizes the results through volcano plots, highlighting the markers with the strongest genetic correlations per SMD. Overall, a clear pattern distinguished MDD from BIP and SCZ; MDD displayed similar genetic correlations across the markers as T2D, CAD and BMI, while BIP and SCZ showed strikingly opposite directions of correlations to MDD, see **Figure 1b**. Accordingly, there was a high negative Spearman’s rank correlation between the LDSC estimates for MDD versus BIP (rs=-.74) and between MDD and SCZ (r_s_=-.83). This contrasts with as well as the positive genetic correlations that exists between these disorders (MDD-BIP r_g_=.44 and MDD-SCZ r_g_=.34; all p<1*10^−16^), see **Figure 1c**. These results suggest that the metabolic pathways involved in MDD etiology differ from those in BIP and SCZ, and that the genetics of metabolic processes may help distinguish between these disorders. Full results are listed in Supplementary Table 2.

**Figure 1.**
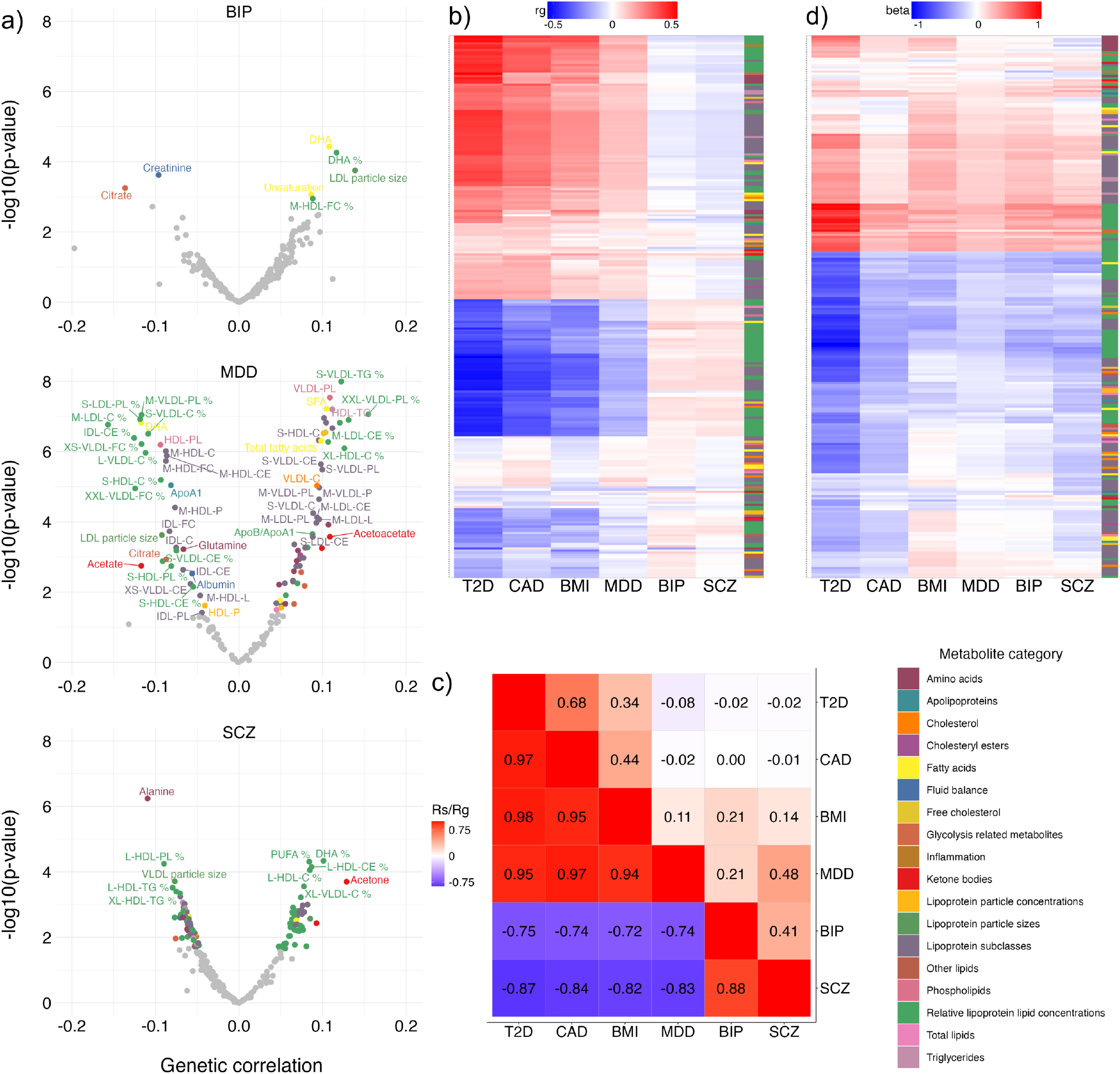
Associations with metabolic markers across the traits. **a)** Using linkage disequilibrium score regression (LDSC), we found widespread significant genetic correlations between the markers and each of the three disorders. These are here summarized through volcano plots, with genetic correlations on the x-axis and -log10(p-values) on the y-axis, separately for the disorders. Significant correlations are indicated in color, corresponding to marker category, and annotated with the marker name. **b)** This panel summarizes the genetic correlations through a heatmap with positive coefficients displayed in red and negative in blue (see legend), for each of the three disorders as well as the additional cardiometabolic traits on the x-axis. The y-axis corresponds to the markers and is sorted based on hierarchical clustering. The column to the right indicates the marker categories as listed in the legend. **c)** A correlation matrix contrasting the genetic correlations between the traits of interest in the upper triangle to the observed Spearman’s correlations of LDSC estimates between these traits across all metabolic markers (as depicted in Figure 1b) in the lower triangle. **d)** Using regression, we also found widespread phenotypic associations between the metabolic markers (y-axis) and with severe mental disorders and cardiometabolic traits and diseases (x-axis); The y-axis is sorted based on hierarchical clustering independently from panel b). MDD=major depressive disorder; T2D=type 2 diabetes; CAD=coronary artery disease; BMI=body mass index; BIP=bipolar disorder; SCZ=schizophrenia.

As a sensitivity analysis, we ran the genetic correlation analyses with the GWAS of metabolic markers corrected for BMI. This yielded the same overall pattern of results, albeit with attenuated strength of correlations, with the LDSC estimates between MDD and the markers still opposite to those for BIP (r_s_=-.57) and SCZ (r_s_=-.43), indicating that the relation between the markers and BMI does not fully explain the divergent patterns between the SMDs.

We further mapped phenotypic relationships between the markers and traits, to compare this to the observed genetic overlap patterns. For this, we used data on plasma concentrations of the Nightingale NMR panel from 207 thousand participants of the UK Biobank. We combined this metabolomics data with information on ICD10 diagnoses for BIP, MDD, SCZ, T2D and CAD, as well as BMI. We estimated phenotypic associations by regressing the traits of interest onto the metabolic markers, covarying for age and sex. The resulting coefficients, displayed in **Figure 1d**, confirmed widespread, highly significant associations between nearly all markers and the traits, with highly similar patterns across all traits (all Spearman’s rank correlations >.76), in line with literature and contrasting with the genetic patterns for BIP and SCZ shown in **Figure 1b**.

Mixed effect directions of genetic variants between pairs of complex traits (concordance), together with differences in polygenicity, may obscure the full extent of their genetic overlap as indicated by global genetic correlations.^22^ We therefore conducted bivariate Gaussian mixture modeling with MiXeR^19^ to estimate the number of shared causal variants between the metabolic markers and the SMDs, irrespective of effect directions. Indeed, MiXeR revealed more extensive overlap than what was indicated by genetic correlations estimated with LDSC, clearly indicating the relevance of metabolic markers for understanding the etiology of these disorders. For example, while BIP had a median absolute genetic correlation of 0.04 with the markers, it shared a median of 85% of their causal variants. Wilcoxon’s signed rank tests indicated highly significant differences (all p<1*10^−12^) of MDD with SCZ and BIP with respect to the distributions of the measures of genetic overlap with the markers, as visualized in **Figure 2a**. Whereas MDD had the highest genetic correlations, BIP shared on average the largest proportion of variants with the markers. This is explained by the concordance rate, which on average was close to 0.54 for BIP, i.e. half of the shared genetic variants have opposing directions of effect, as opposed to 0.79 for MDD and 0.41 for SCZ. **Figure 2b** provides an example of this by illustrating the genetic overlap between SMDs and glycoprotein acetyls. This shows that, whereas the number of shared variants is similar for all three disorders, MDD has a much higher concordance rate, explaining the higher genetic correlation, and suggesting a more direct genetic relation with metabolic markers than BIP and SCZ. The overlap and concordance estimates for each pair of disorder-marker are provided in Supplementary Table 3.

**Figure 2.**
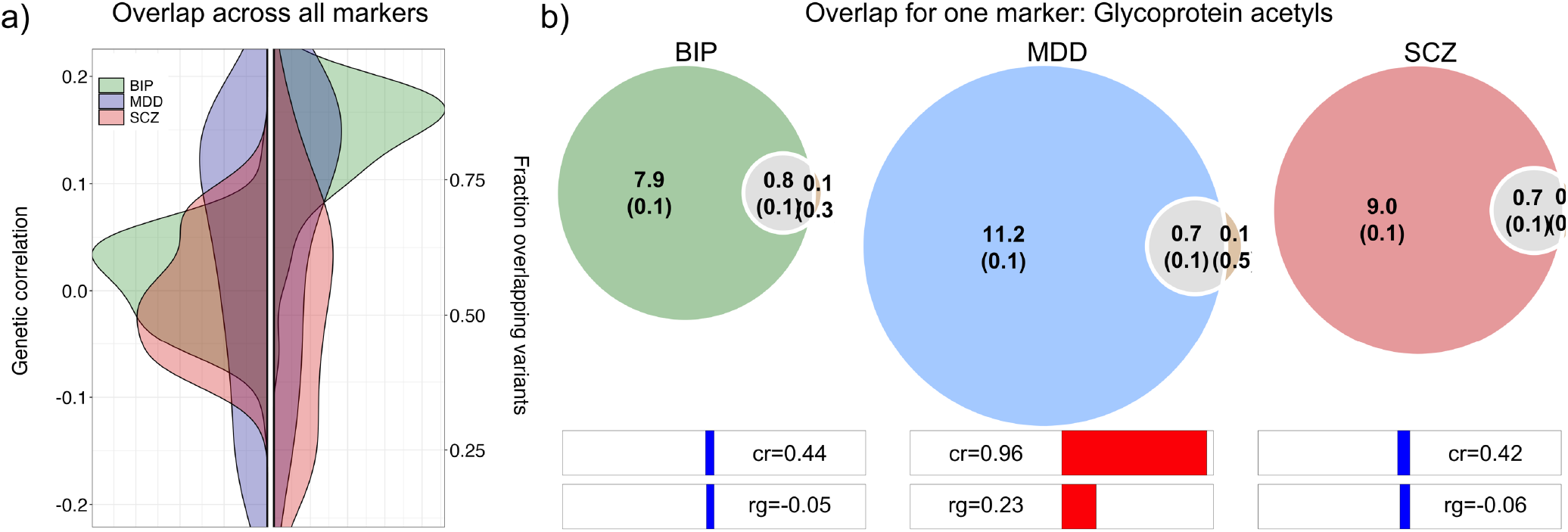
Complementary measures of genetic overlap of the metabolic markers with the severe mental disorders (SMDs). **a)** Density plots showing the distributions of genetic overlap measures observed across all of the 249 markers and the three SMDs (colors, see legend). This summarizes the genetic correlations (left) and fraction of the markers’ causal variants shared (right), as indicated on the y-axis. **b)** Venn diagrams illustrating the genetic overlap between the three disorders and glycoprotein acetyls, as an example for one marker. The estimated number of unique and shared variants are indicated in thousands, with standard deviations in brackets. Below the Venn diagrams are bars indicating the concordance rates (‘cr’) of effect directions of shared variants, whereby 0.50 would mean an equal number of variants with opposing and same directions of effects on the pair of traits. The bar at the bottom indicates the estimated genetic correlation (‘rg’) of the pairs. BIP=bipolar disorder; MDD=major depressive disorder; SCZ=schizophrenia.

### Causal relationships

We conducted bidirectional Mendelian randomization (MR) to identify significant causal relationships between the markers and the SMDs. **Figure 3** lists the inverse variance weighted (IVW) MR coefficients that were significant after correction for multiple comparisons, which were also significant through weighted median and MR Egger approaches, confirming the robustness of the findings. As can be seen in **Figure 3a**, there were causal effects of numerous markers on each of the three disorders, with both shared and trait-specific effects. The findings in the opposite direction (**Figure 3b**) reiterated the distinction between MDD on the one hand and BIP and SCZ on the other; only MDD shows widespread significant causal influences on the markers.

**Figure 3.**
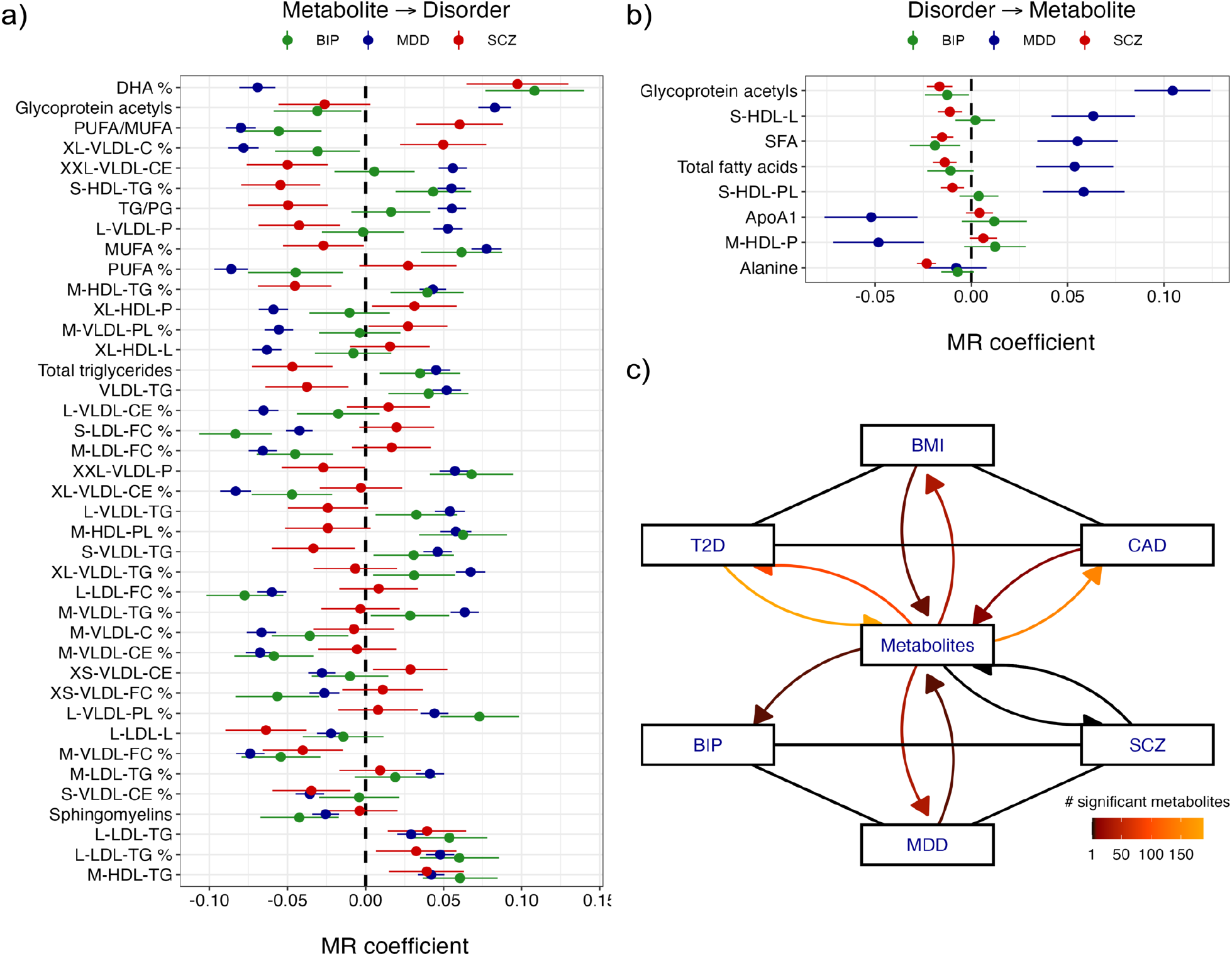
Bidirectional causal relationships between the metabolic markers and the traits. **a)** The causal effects of the significant markers on the severe mental disorders (SMDs), with inverse variance weighted Mendelian randomization (MR) coefficients (dots) and their standard errors (lines) displayed on the x-axis, the markers listed on the y-axis, and the three severe mental disorders (SMDs) represented by colors as indicated in the legend. **b)** same as panel a) but in opposite direction, i.e. the significant causal effects of the disorders on the markers. For both panels, the markers are ordered on the y-axis by Cochran’s Q-statistic, which captures the extent of divergence of the MR coefficients between the three disorders; the markers which show the most divergent relationship between the three disorders are at the top. **c)** Pathways of causal relationships between all traits of interest. Connections indicate robust significant causal effects as identified by all three MR approaches (see text). The curved arrows connect the traits to the markers, color coded by the number of significant markers. BIP=bipolar disorder; MDD=major depressive disorder; SCZ=schizophrenia; BMI=body mass index; T2D=type 2 diabetes; CAD=coronary artery disease.

We subsequently expanded the MR analyses to investigate BMI (a prominent risk factor) and CAD and T2D (common comorbid conditions) as potentially clinically relevant mediators of the relationships between the SMDs and metabolic markers. We found no robust evidence of direct causal effects of these additional traits on the SMDs or *vice versa*, despite their well-known phenotypic relationships. Rather, the estimated causal relationships indicated that the metabolic markers are intermediate. The findings, summarized in **Figure 3c**, indicated that BMI increases the risk of T2D and CAD, while all three have bidirectional causal relationships with the markers. As can also be deduced from **Figure 3a** and **b**, MDD had a substantially higher number of bidirectional causal relationships with the markers than BIP and SCZ. Overall, more metabolic markers influenced the SMDs and cardiometabolic traits than the other way around, indicating their potential as modifiable factors that may be targeted to treat the disorders and their cardiometabolic comorbidities. Full results, across all pairs, are provided in Supplementary Tables 4-6.

### Molecular mechanisms

To further delve into the biological pathways coupling the SMDs to metabolic health, we ran conjunctional false discovery rate (cFDR) analyses.^23^ Through cFDR, we identified hundreds of genetic variants shared by each of the 3 × 249 disorder-marker pairs, listed in Supplementary Tables 7-9. The shared lead variants were then mapped to genes using OpenTargets,^24^ which in turn were tested for enrichment among Gene Ontology (GO) biological processes. **Figure 4a** shows how much the aggregate of genes identified through the conjunction of the disorders and markers is shared across, or specific to, the three disorders. **Figure 4b** lists the top 5 most significant GO terms for each disorder among the 249 marker pairings. Overall, identified pathways were relevant for energy metabolism or neuronal processes. For the BIP-marker pairs, general metabolic and mitochondrial pathways were common among the list of significant pathways; for MDD, the overlap with markers related most to (organ) developmental processes in addition to many pre-and postsynaptic processes; for SCZ, besides these same processes, we mostly found pathways related to responses to biotic stimuli and the immune system. Thus, the disorders also have shared as well as specific biological processes that mediate their relationship with metabolic markers. Supplementary Tables 10-12 provide a comprehensive overview of all significant GO terms and their enrichment, per disorder.

**Figure 4.**
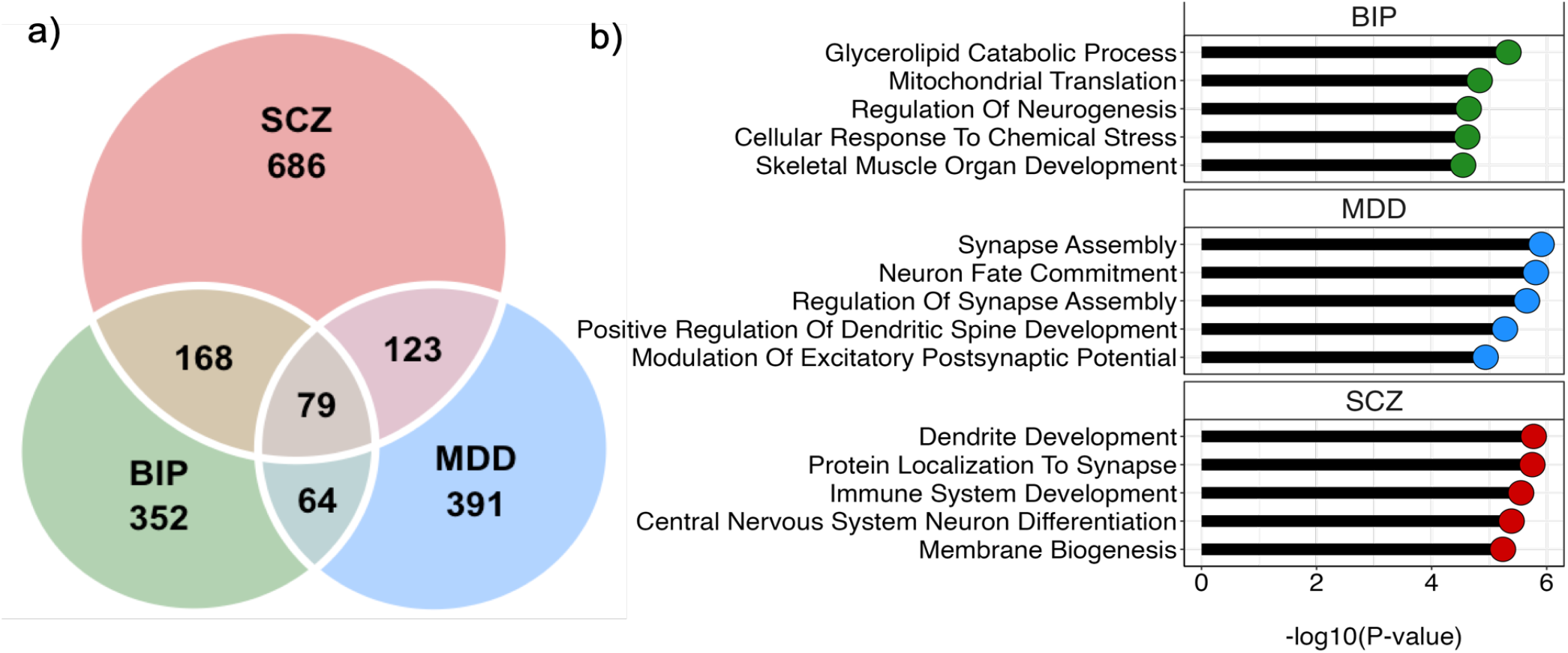
Coupling of genes overlapping between severe mental disorders and metabolic markers to gene ontology terms. **a)** Venn diagram depicting the number of unique genes found through the conjunctional analyses, aggregated across all markers, for each disorder. **b)** Plots showing the top 5 most significantly enriched gene ontology terms (y-axis) based on the mapped genes, per disorder (panels), with the observed p-values (x-axis). BIP=bipolar disorder; MDD=major depressive disorder; SCZ=schizophrenia

In order to gain an overview of the relevance of the findings for brain- and bodily health, we coupled the three aggregated sets of mapped genes found through the cFDR approach to tissue gene expression data. For all three disorders, there was significant enrichment across multiple brain regions and body tissues, without a clear distinguishing pattern between the disorders. **Figure 5a** shows regional variation in expression across cortical and subcortical brain regions, with the most significant region being the superior temporal sulcus, for SCZ (p=9.5×10^−11^). **Figure 5b** shows the enrichment, per GTEx v8 general body tissue type. This indicates that the mapped genes are not expressed solely in the brain, as the heart and liver were also among the most implicated organs.

**Figure 5.**
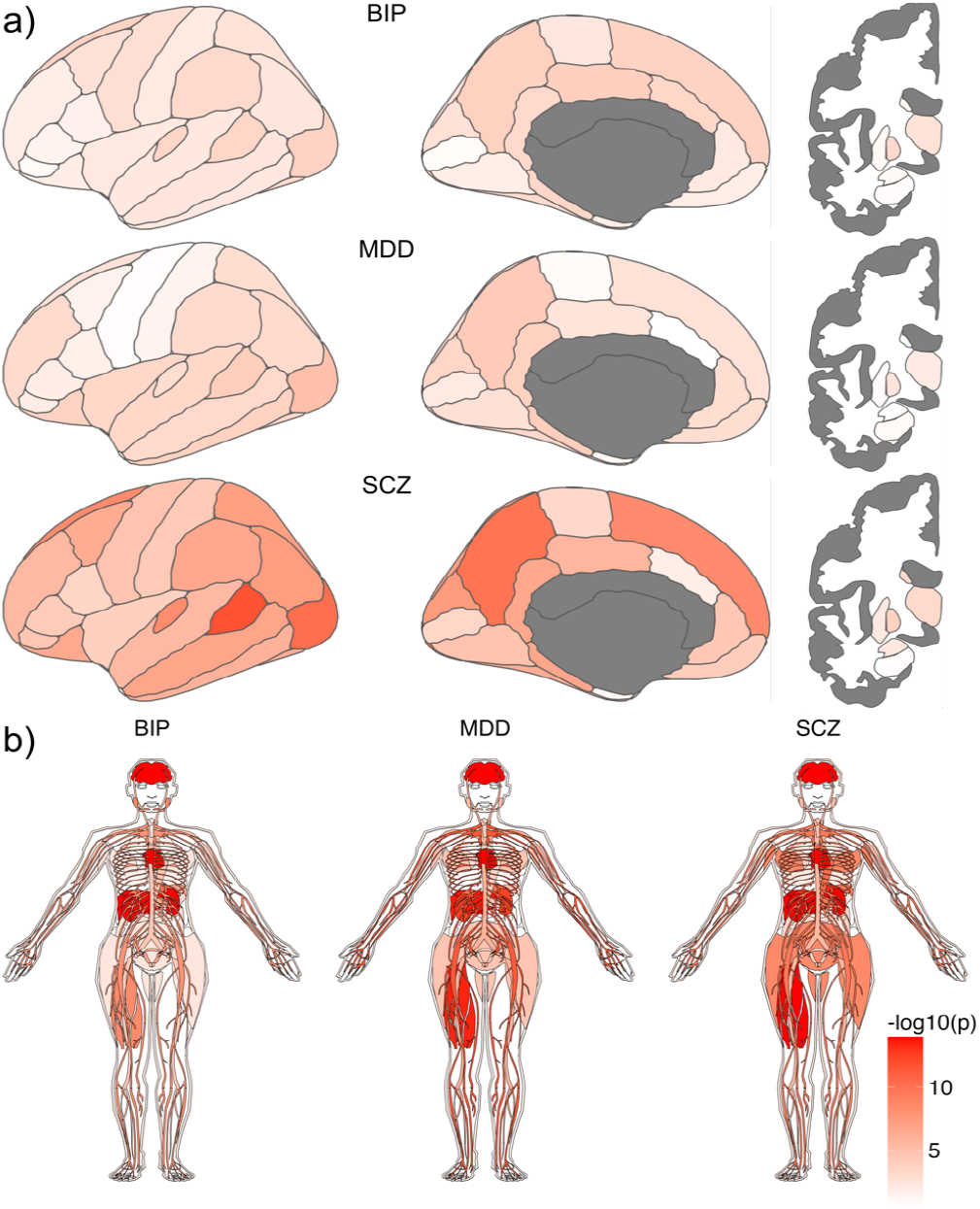
Expression patterns of genes shared between the metabolic markers and disorders, across body and brain tissue. Color coding indicates significance as -log10(p-value) of tests checking enrichment of the lists of shared genes, per disorder, among expressed genes. **a)** Brain maps summarizing significance of enrichment tests across cortical (Desikan-Killiany atlas) and subcortical (Freesurfer aseg) regions, based on expression data from the Allen brain atlas. **b)** Anatograms visualizing the results of the enrichment tests for each of 30 general tissue types from the GTEx database. BIP=bipolar disorder; MDD=major depressive disorder; SCZ=schizophrenia.

## Discussion

Here, we performed large-scale analyses that revealed extensive genetic overlap of SMDs and related cardiometabolic traits with metabolic markers. Most notably, we found differences in the shared genetic architecture of these markers with MDD on the one hand and BIP and SCZ on the other, which contrasted with their phenotypic associations. Follow-up analyses confirmed extensive causal effects of the metabolic markers on the SMDs, while MDD also showed effects on the markers. Functional annotation of genetic variants shared by the SMDs and metabolic markers led to the identification of specific biological processes relevant for metabolism and neuronal activity. Gene expression across regional brain and body tissues showed a distributed effect of shared genetic variants. Taken together, the current findings underscore the role of metabolic processes in SMDs, and highlight the potential of metabolic psychiatry.

SCZ, BIP and MDD have substantial genetic and clinical overlap,^25,26^ yet we found their genetic relationship with the metabolic markers to be distinctly divergent from their phenotypic overlap. The observed patterns of genetic overlap with metabolic markers suggest that the genetic underpinnings of MDD are more similar to cardiometabolic diseases than those of SCZ and BIP. The similarity in cardiometabolic comorbidities across these SMDs,^1,27^ opposing the observed patterns of genetic correlation, may therefore arise from environmental influences on SCZ and BIP. This is likely to encompass an unhealthy lifestyle, including poor diet and sedentary behaviour,^28,29^ as well as obesogenic effects of psychotropic medication.^30^ Our findings are in line with previous reports of SCZ and BIP being genetically associated with lower BMI,^31,32^ while MDD is genetically associated with higher BMI.^31^ Taken together, they indicate that factors beyond common genetic variants play a significant role in obesity and other cardiometabolic comorbidities in SCZ and BIP, whereas in MDD, genetic and non-genetic factors appear to act in the same direction. Characterization of the role of confounding and modifiable risk factors will be of high value to disentangle disease-specific etiologies and differential diagnosis, including subtyping.^33^ This is important for the development of targeted interventions, as well as individualized prediction of disease onset and clinical outcomes. Our findings support previous claims that metabolomics data may play a central role in such efforts.^12^

We found robust evidence of causal relationships between metabolic markers and the SMDs, confirming and expanding on previous findings,^34,35^ underlining the potential of these markers as targets for interventions. Stronger bidirectional effects were observed for MDD compared to SCZ and BIP, in line with higher genetic correlations for MDD. The results indicate that genetic variants influencing both MDD and metabolic markers work in concert with non-genetic factors, while the etiology of SCZ and BIP involve more mixed patterns of directions. The three metabolic markers with the strongest causal influence had divergent, opposite directions of effects on the three SMDs. Docosahexaenoic acid (DHA), a polyunsaturated fatty acid (PUFA) essential for synaptic membrane function, memory and neuroprotection,^36,37^ has previously been identified as having a causal influence on MDD.^38^ The levels of DHA and other PUFAs, and their ratio to monounsaturated fatty acids (MUFAs), have also been found to be different between the three SMDs in postmortem brain tissue.^39,40^ Additionally, glycoprotein acetyls, an inflammatory marker and early indicator of cardiovascular risk,^41^ has been associated with depression in large clinical cohorts^42^ and bipolar depressive episodes.^43^ The identification of causal effects of these markers may be leveraged to better understand how metabolic pathways have divergent effects on the three SMDs. We found no robust direct causal relationships between the SMDs and the cardiometabolic traits, suggesting their associations are primarily downstream of other factors, including those that influence metabolic markers. Our findings thereby suggest metabolic pathways can play a substantial role in explaining multimorbidity, as previously reported across common noncommunicable diseases.^44^

The hundreds of identified genetic variants that influence both the SMDs and the markers were mapped to genes known to be involved in specific biological processes of relevance for energy metabolism and neurotransmission, aligning with literature on the role of metabolic pathways in these disorders.^45^ While previous genetics studies of these SMDs have emphasized a near-complete overlap in causal variants,^19^ our approach did identify a substantial unique set of genes and pathways for each disorder. Consequently, despite the high global overlap between SMDs, our findings reveal unique pathways specific to each disorder that are linked to metabolism, enabling new opportunities for tailored drug discovery and treatment of each disorder. As one example, our findings indicated a role for mitochondrial dysfunction in BIP, fitting with the known mode of action of lithium treatment.^46^ The importance of metabolic pathways for treatment is further highlighted by the recent line of findings on GLP-1 receptor agonists as treatment for SMDs.^47^ Coupling the overlapping genes to expression data across regional brain and body tissues highlighted organs central to cardiometabolic function, such as the liver and heart, as well as the brain, consistent with research on the overlap between body and brain measures^48^ and their relevance for SMDs.^49–51^ The involvement of these organs is further in line with the substantial somatic comorbidities of these disorders, emphasizing the importance of an integrated approach focusing on the brain and the body to understand and treat SMDs.^52^

Together, these findings implicate metabolic pathways in SMDs’ etiology. We found divergent patterns of genetic correlations with metabolic markers across SMDs, with important yet heterogeneous link to cardiometabolic comorbidities. These findings suggest that metabolomic data has potential for guiding the development of targeted interventions, and emphasize the importance of metabolic psychiatry research to better understand the etiology of SMDs.

## Data Availability

The data incorporated in this work were gathered from public resources. The code is available via https://github.com/precimed (GPLv3 license). Correspondence and requests for materials should be addressed to d.v.d.meer[at]medisin.uio.no

## Methods

### Phenotypic associations

For the UKB, we obtained data under accession number 27412. The composition, set-up, and data gathering protocols of the UKB have been extensively described elsewhere.^53^ It has received ethics approval from the National Health Service National Research Ethics Service (ref 11/NW/0382), and obtained informed consent from its participants. For the primary analyses, we selected unrelated White Europeans (KING cut-off 0.05)^54^ that had the Nightingale NMR metabolomics data, as well as complete covariate data available (N=207 836, mean age 57.4 years (SD=8.0), 53.7 % female).

We applied additional pre-processing through the ‘ukbnmr’ R package to the NMR data as released by UKB, to remove sources of technical noise.<u>^55^</u> We further applied rank-based inverse normal transformation,<u>^56^</u> leading to normally distributed measures.

ICD10 diagnoses for BIP (F31, N=769), MDD (F32, N=13,887), SCZ (F20, N=447), T2D (E11, N=17,299) and CAD (I20-I25, N=25,638) were taken from UKB field 42170. BMI was taken from UKB field 21001, with a mean of 27.4 (SD=4.8).

We ran a series of logistic regressions for the diagnoses (dummy-coded), regressing these onto each individual marker, covarying for age and sex, and recorded the log odds coefficient from these models. We z-scaled BMI, as a continuous measure, and recorded the resulting linear regression coefficient.

### GWAS summary statistics and pre-processing

We included GWAS summary statistics of all 249 metabolic markers from the Nightingale NMR metabolomics panel.<u>^11^</u> We previously generated these summary statistics, based on 207,841 participants from the UK Biobank and 92,645 participants from the Estonian Biobank, meta-analyzed together.^18^ For SCZ,^13^ MDD,^14^ BIP,^15^ CAD,^16^ T2D^17^ and BMI^20^ we made use of the latest European-specific GWAS summary statistics. Where available, we selected versions of the summary statistics that did not contain UKB or EstBB data, in order to minimize sample overlap as this may create biases for the MR and cFDR analyses. All summary statistics were QC’ed and formatted using our standardized pipeline (https://github.com/precimed/python_convert/blob/master/sumstats.py).

### Bivariate LDSC

We applied cross-trait^21^ LDSC to estimate genetic correlations between the metabolic markers, SMDs and cardiometabolic traits. For this, we formatted the GWAS summary statistics in accordance with recommendations, including ‘munging’ and removal of all variants in the extended major histocompatibility complex (MHC) region (chr6:25–35 Mb).

### Bivariate MiXeR

We applied a bivariate Gaussian mixture model, MiXeR,^19,57^ to estimate the number of shared causal genetic variants between the three SMDs and each metabolic marker, for a total of 3 * 249 analyses. MiXeR models additive genetic effects, based on the GWAS summary statistics, as a mixture of four components, representing null SNPs in both traits (π_0_); SNPs with a specific effect on the first and on the second trait (π_1_ and π_2_, respectively); and SNPs with non-zero effect on both traits (π_12_). For the mathematical framework, please see a previous publication.^19,57^ We checked whether the models, for each pair of traits, generated a positive AIC value, compared to a baseline infinitesimal model, indicating good model fit. All models passed this quality check.

### Mendelian randomization

We ran bidirectional MR, investigating the causal relationships between the SMDs, the 249 metabolic markers and the cardiometabolic traits. For this, we applied the TwoSampleMR R package to the GWAS summary statistics. We selected only genome wide significant variants for the analysis, clumped using PLINK with clump_p = 1, clump_r2 = 0.001, clump_kb = 10000 against the 1000 Genomes Phase3 503 EUR samples keeping other settings default. We calculated MR regression coefficients using the IVW method. To ensure robust findings, we further applied the weighted median method^58^ and the MR-Egger method.^59^ We only selected findings that showed an FDR multiple comparisons-significance for both the IVW and weighed median approach, together with nominal significance for the MR Egger method.

### Conjunctional FDR

We conducted conjunctional false discovery rate (cFDR) analyses for each of the three SMDs with each of the 249 markers, i.e. 3 × 249 analyses. The cFDR analyses consisted of conditioning the GWAS summary statistics for each of the pair of traits onto each other in both directions through the pleioFDR tool using default settings, see https://github.com/precimed/pleiofdr, whereby the highest of the resulting pair of FDR values per genetic variant was taken as the strength of evidence for its shared effects. We set an FDR threshold of 0.05 as whole-genome significance, in accordance with recommendations.

### Gene mapping

We used the Variant-to-Gene (V2G) pipeline from Open Targets Genetics, to map lead variants from the cFDR analyses to genes based on the strongest evidence from quantitative trait loci (QTL) experiments, chromatin interaction experiments, in silico functional prediction, and proximity of each variant to the canonical transcription start site of genes.^24^

### Gene set enrichment

We carried out gene set enrichment analyses on the mapped genes, investigating terms that are part of the Gene Ontology biological processes subset (n=7522), as listed in the Molecular Signatures Database (MsigdB; c5.bp.v7.1). We ran Fisher exact tests on the mapped genes from each individual cFDR analysis (i.e. 249 × 3 sets of mapped genes), leveraging the enricher R package. We restricted the output to those terms that had more than 5 overlapping genes and that were smaller than 500 genes in total, with a q-value <.05. We further removed terms from the list of significant enrichments if more than 80% of their overlapping genes were among more significant terms. Running the enrichment analyses on individual cFDR sets of genes, with the above settings, was chosen to ensure that we identified more biologically specific terms than when running across aggregated gene lists and included larger pathways.

### Tissue expression

We used Fisher exact tests to calculate the significance of overlap between the three sets of cFDR mapped genes (aggregated within disorder across cFDR runs) and predetermined sets of differentially expressed genes (DEG) in each of the 30 general tissues available in the GTEXv8 database. DEGs are defined as genes with log2 transformed, normalized expression values (Read Per Kilobase per Million, zero-mean) with P-value ≤ 0.05 after Bonferroni correction and absolute log fold change ≥ 0.58 in a given tissue.

For the brain-based expression analyses, mRNA distribution data was acquired from the Allen Human Brain Atlas.^60^ In cases where multiple probes were available for a specific mRNA, the probe with the highest differential stability was selected.^61^ For the current study, we summarized the data using a Desikan-Killiany atlas-based map with the Python toolbox Abagen.^62^ Data normalization was performed using the default scaled robust sigmoid method.^61^ To assess the difference in gene expression between the list of identified genes and all other genes, a Wilcoxon rank-sum test was conducted for each brain region.

### Statistical analyses

All pre-processing steps and analyses performed outside the above-mentioned tools and software, e.g. formatting the data and creating the graphs, were carried out in R, v4.2.

## Acknowledgements

This work was partly performed on the TSD (Tjeneste for Sensitive Data) facilities, owned by the University of Oslo, operated and developed by the TSD service group at the University of Oslo, IT-Department (USIT). Computations were also performed on resources provided by UNINETT Sigma2 -the National Infrastructure for High-Performance Computing and Data Storage in Norway. Computation of Estonian Biobank data was carried out in part in the High-Performance Computing Center of University of Tartu. The Estonian Biobank Research Team, responsible for data collection, genotyping, QC, and imputation, comprises Andres Metspalu, Tõnu Esko, Reedik Mägi, Mari Nelis and Georgi Hudjashov.

## Materials & Correspondence

The data incorporated in this work were gathered from public resources. All data described are available through UK Biobank, subject to approval from the UK Biobank access committee. See https://www.ukbiobank.ac.uk/enable-your-research/apply-for-access for further details. The MOSTest code is available via https://github.com/precimed/mostest (GPLv3 license). GWAS summary statistics are uploaded to the NHGRI-EBI GWAS catalog (https://www.ebi.ac.uk/gwas/).

## Author contributions

D.v.d.M. and O.A.A. conceived the study; D.v.d.M., Z.R., A.O., and A.S. pre-processed the data. D.v.d.M. performed all analyses, with conceptual input from S.S., J.R., A.S. and O.A.A.; All authors contributed to interpretation of results; D.v.d.M. drafted the manuscript and all authors contributed to and approved the final manuscript.

